# Neuroimaging-derived brain endophenotypes link molecular mechanisms to Alzheimer’s disease and aging

**DOI:** 10.1101/2025.11.25.25340884

**Authors:** Ruixi Li, Ru Feng, Anjing Liu, Xuewei Cao, Philip L. De Jager, David Bennett, The Alzheimer’s Disease Functional Genomics Consortium, Christos Davatzikos, Junhao Wen, Gao Wang

## Abstract

Alzheimer’s disease (AD) genome-wide association studies (GWAS), typically based on clinical phenotypes, have identified numerous risk loci, yet linking these variants to brain changes and molecular processes remains challenging. We developed a DNE-xQTL framework integrating deep learning-derived dimensional neuroimaging endophenotypes (DNEs) with comprehensive brain molecular quantitative trait loci (xQTL) to dissect genetic pathways underlying AD– and aging-related brain variation. By performing GWAS on seven DNEs and applying integrative computational analyses, we biologically annotated each DNE and prioritized xQTL-supported gene targets. This approach both enhanced interpretation of established AD loci through DNE-mediated annotations and revealed underexplored regulatory pathways, organizing 209 candidate genes into evidence-based tiers. We highlight three regulatory clusters: glutamate-receptor and mitochondrial pathways implicating excitatory-neuron vulnerability, SREBP2-associated cholesterol homeostasis linked to vascular dysfunction, and primary-cilia-associated transport implicated in aging. By connecting pre-symptomatic brain alterations to molecular targets and relevant cell types, this framework may inform earlier risk stratification before clinical neurodegeneration occurs.

## INTRODUCTION

Understanding the molecular architecture of pre-symptomatic brain aging is critical for identifying therapeutic targets before irreversible neurodegeneration occurs. Traditional endophenotypes, including cerebrospinal fluid (CSF) biomarkers (amyloid-β, tau), PET imaging, and structural MRI measures (hippocampal volume, cortical thickness), typically quantify single features and often miss subtle early changes that vary across individuals. Beyond practical limitations of CSF invasiveness and PET costs and radiation exposure, these univariate measures fundamentally fail to capture the heterogeneous spatial patterns characteristic of individual aging trajectories and disease presentations. Dimensional neuroimaging endophenotypes (DNEs), derived from deep learning applied to multivariate MRI data, address these limitations by extracting complex brain-wide patterns as quantitative dimensions that capture individual variability and detect subtle pre-symptomatic alterations. However, DNEs remain abstract imaging constructs lacking biological interpretability: the cellular processes driving these brain changes are uncharacterized, limiting molecular target discovery for early intervention.

Genome-wide association studies (GWAS) have identified approximately 100 AD risk loci but cannot reveal how genetic variants manifest as brain changes or disease mechanisms^1,2^. Limitations in AD phenotyping further complicate gene discovery. Clinical AD diagnosis demonstrates limited accuracy, with autopsy studies revealing 10–30% discordance due to phenotypic overlap with vascular, Lewy body, and other pathologies^3^; moreover, recent large-scale GWAS increasingly rely on proxy phenotypes from parental history reports^4,5^, which introduce survival bias and nonrandom participation effects that produce misleading genetic correlations^6^. Endophenotypes, as intermediate traits, offer a path forward by enabling genetic variants to serve as instrumental variables for causal inference^7^. Recent studies leveraged this principle, performing GWAS on DNE dimensions followed by Mendelian randomization (MR) against disease GWAS, establishing putative causal relationships from DNE to AD risk^1,2^. While demonstrating that imaging patterns influence endpoints, this approach cannot explain underlying molecular mechanisms or identify therapeutic targets. Separately, molecular quantitative trait loci (xQTL) mapping links genetic variants to molecular phenotypes (e.g. gene expression and protein levels), revealing how variation influences brain molecular states but not how these connect to *in vivo* pre-symptomatic brain alterations. Each approach addresses only part of the pathway. DNE-endpoint integration may establish causality but remains mechanistically opaque; xQTL-endpoint integration identifies molecular associations but lacks intermediate phenotypes to dissect how molecular changes manifest as observable brain alterations. No study has systematically integrated imaging endophenotypes, comprehensive multi-context brain xQTLs, and endpoint GWAS to simultaneously characterize what DNEs represent biologically and connect them to neurodegeneration.

Here, we developed an integrative genetic framework applied to seven MRI– and AI-derived DNEs from the MULTI and iSTAGING consortia (n=32,000–43,100), large-scale AD and aging GWAS (n=462,666–1,958,774), and brain xQTL from the FunGen-xQTL consortium across 48 molecular contexts from 2,328 individuals (**Figure 1**) to systematically map the molecular architecture of pre-symptomatic brain aging and neurodegeneration. This *DNE-xQTL framework* enabled multi-scale evidence synthesis through fine-mapping and multi-GWAS colocalization to identify putative causal genetic loci; brain xQTL mapping to link variants to genes across molecular modalities and cell types; comprehensive transcriptome-wide association studies (TWAS) to establish gene-level evidence; integration of variant-level colocalization with gene-level TWAS to reduce false-positive gene identification, with cell-type-resolved causal analyses applied to genomically complex loci containing multiple candidates; and pathway analysis to biologically annotate each DNE. We revealed that each DNE captures distinct cellular processes (*iAD1*: cytoskeletal dysfunction; *iAD2*: excitotoxicity; *iAging1-5*: lipoprotein transport, synaptic protein degradation, autophagy-extracellular vesicle impairment, mitochondrial adaptation, and neuroinflammation-driven vascular damage). By consolidating loci-level and gene-level evidence linking DNEs to AD and aging (telomere length), we organized 209 genes into evidence-based tiers and identified three gene clusters that may connect DNEs to clinical outcomes: a synaptic dysfunction cluster revealing oligodendrocyte contributions to glutamate receptor trafficking and excitotoxicity (*TNIP1*, *NSF*, *SEZ6L2*); a vascular-cholesterol cluster linking cholesterol dysregulation to cerebrovascular integrity (*NBEAL1*, *SCAP*); and a primary cilia cluster connecting AD to aging through intraflagellar transport and extracellular vesicle pathways (*CBY1*, *MAP4*, *BIN1*). This hypothesis-generating framework enables biologically motivated gene prioritization beyond what conventional GWAS and xQTL studies can provide, revealing candidate molecular targets for early intervention.

**Figure 1.**
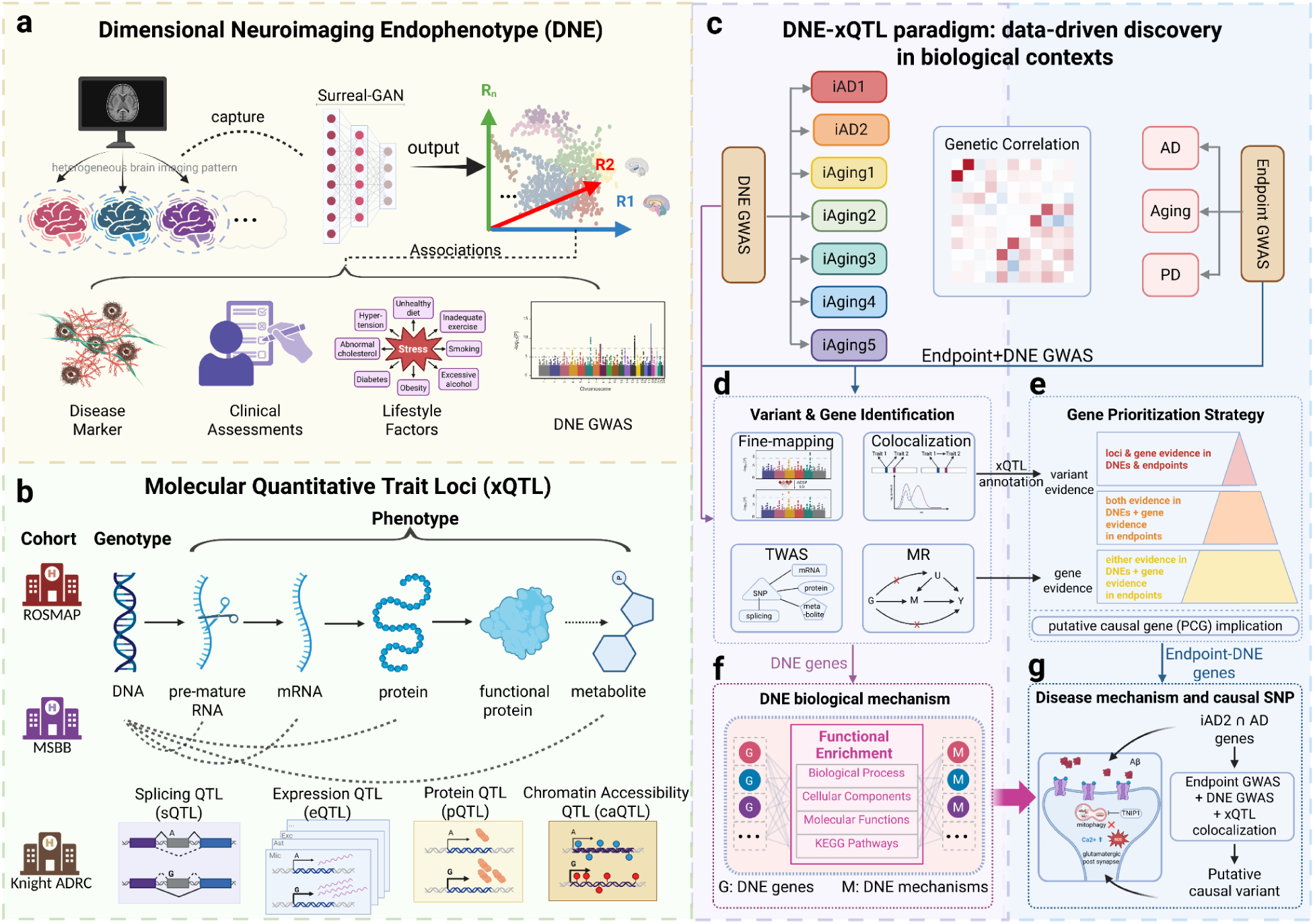
The DNE–xQTL analytical framework bridges genetics, neuroimaging, and molecular mechanisms to reveal disease biology. **(a)** DNE generation from brain MRI across up to 43,100 individuals using deep learning trained on AD and aging outcomes. **(b)** Comprehensive brain molecular QTL data across multiple modalities (eQTL, sQTL, pQTL, etc.) and diverse cell types from the FunGen-AD consortium. **(c)** Genetic correlations across 16 GWAS traits to assess polygenic similarity and heritability enrichments to pinpoint DNE contributions to endpoints. **(d)** Variant and gene identification: fine-mapping and multi-GWAS colocalization identify putative causal variants; brain xQTL mapping across molecular contexts links variants to regulated genes; TWAS and Mendelian Randomization (MR) identify putative causal genes. (e) Gene prioritization strategy: genes supported by both variant-level and gene-level evidence are organized into tiers, with convergent evidence ranked highest; putative causal gene (PCG) implication addresses loci containing multiple candidate genes. **(f)** Functional annotation via GO/KEGG enrichment assigns biological context for each DNE. **(g)** Integration of DNE biological annotations with colocalization across endpoints, DNEs, and xQTLs identifies putative causal variants linking pre-symptomatic brain changes to disease. This framework enables generating testable hypotheses linking abstract imaging patterns to molecular targets with cellular resolution.

## RESULTS

### Genetic fine-mapping and multi-trait colocalization identify shared architecture between neuroimaging endophenotypes and neurodegeneration

For the seven DNE traits (*iAD1*, *iAD2*, *iAging1*, *iAging2*, *iAging3*, *iAging4*, and *iAging5*), we conducted fine-mapping and multi-GWAS colocalization analyses to pinpoint putative causal variants and assess genetic sharing across traits. *iAD1-2* represents Alzheimer’s disease related imaging dimensions trained on Alzheimer’s Disease Neuroimaging Initiative (ADNI) case and control data. *iAging1-5* consists of different normative aging dimensions derived from a large multi-cohort population dataset from the iSTAGING consortium^1,8^. The trained models were applied to UKBB populations (n=32,829–33,541) to get continuous scores for each DNE trait, followed by GWAS adjusting for age, sex, total intracranial volume (ICV) and genetic principal components (**Methods**). Sample size of DNE identification and GWAS, total count of the overall and significant variants for each DNE were summarized in **Table S1**. Bivariate linkage disequilibrium score regression (LDSC) quantified genome-wide polygenic sharing across DNE pairs (**Figure 2**), with strongest genetic correlations occurred between *iAD2* and *iAging2* (r_g_ = 0.70, P = 1.53×10^−108^), and between *iAD1* and *iAging3* (r_g_ = 0.67, P = 6.17×10^−94^), with additional significant correlations observed among other DNE pairs (**Figure S1**, **Table S3**).

**Figure 2.**
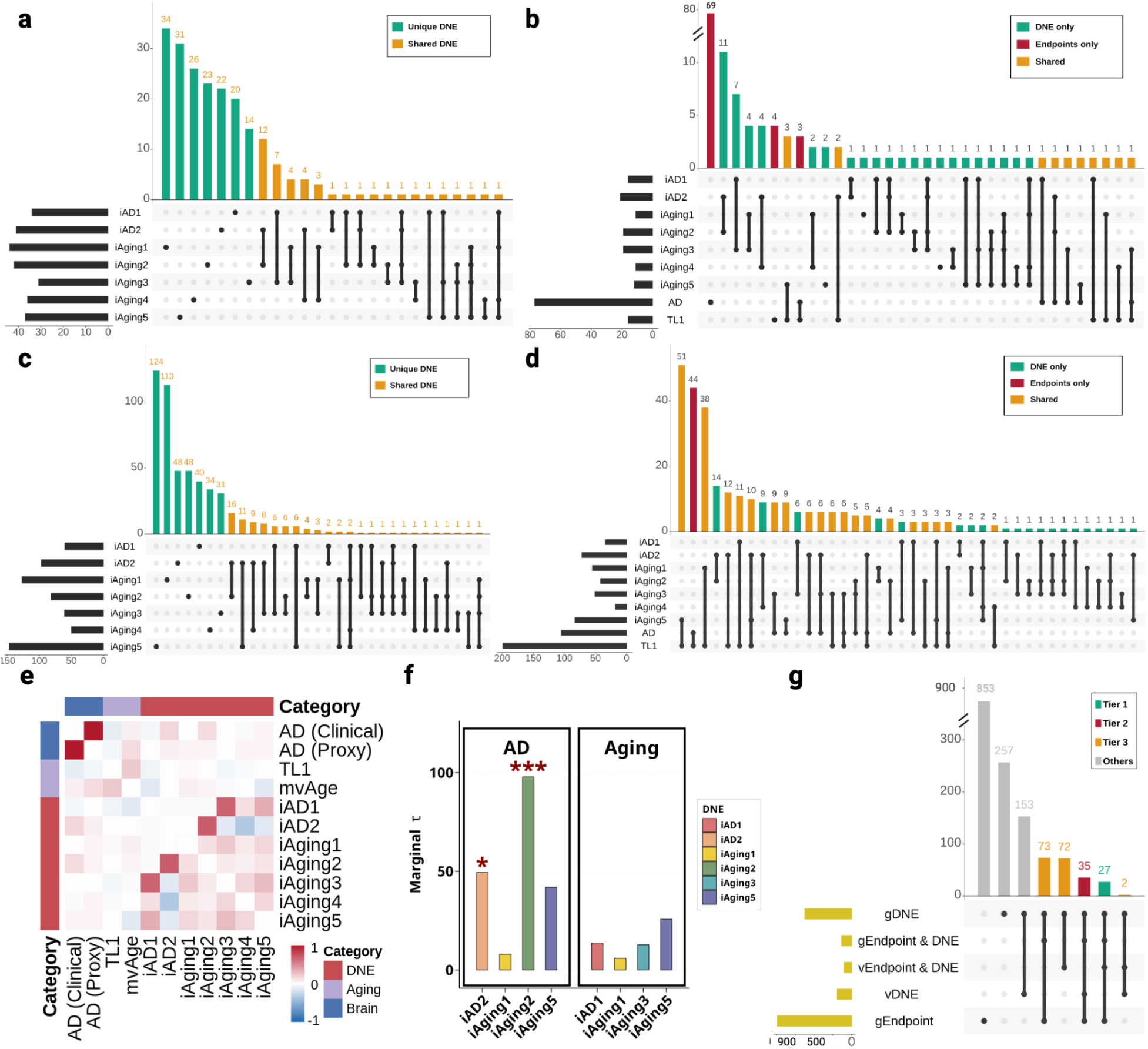
Putative causal variants and genes across DNE, Alzheimer’s disease (AD) and aging. **(a)** Multi-GWAS colocalization identified 95% colocalization sets (CoS) across seven DNEs, showing loci-level patterns of sharing. **(b)** DNE-endpoint colocalization further identifies shared genetic architecture with AD and Aging (telomere length). **(c)** UpSet plot of genes identified by transcriptome-wide association studies (TWAS) integrating brain xQTL across seven DNEs, showing gene-level patterns of sharing. **(d)** DNE-endpoint gene overlaps from TWAS reveal extensive shared genetic architecture: 238 genes with telomere length, genes overlap between DNEs and AD, including 14 genes showing three-way convergence across AD, telomere length, and DNEs. The use of DNE enables fine-grained dissection at both variant-level (A-B) and gene-level (C-D) resolutions. **(e)** Bivariate LDSC across all GWAS used, assessing polygenetic similarity. **(f)** Stratified LD score regression using both maxVCP from colocalization and maxPIP from fine-mapping quantifies heritability enrichment for each DNE in AD and aging, showing positive enrichments with asterisks indicating levels of statistical significance (*iAging2* in AD: τ=0.54, P=3.4×10^−5^; *iAging5* in AD: τ=0.34, P=6.5×10^−3^), establishing DNE relevance as intermediate phenotypes. **(g)** UpSet plot of gene-level evidence across DNE-endpoint pairs with a 3-tier evidence framework organizes candidate genes: Tier 1 (causal variant and gene evidence in endpoints and DNEs); Tier 2 (both evidence types in DNEs, and gene evidence in endpoints); Tier 3 (either evidence in DNEs, and gene evidence in endpoints).

Genome-wide fine-mapping^9^ with a new Linkage Disequilibrium (LD) reference panel for European ancestry^10^ identified 2,131 variants within 83 95% credible sets (CS) across seven DNEs (**Methods**; **Figure S2**). Multi-GWAS colocalization using *ColocBoost*^11^ identified 1,690 variants within 43 95% colocalized confidence sets (CoS) shared across DNE traits (**Figure 2a, Figure S3**). The distribution of variants per credible set (CS) from fine-mapping analysis showed that 93% (40/43) of CS contained fewer than 100 variants, and colocalization analysis revealed that 91% (60/66) of CoS contained fewer than 100 variants, demonstrating that both methods achieve high-resolution genetic mapping for most regions (**Figure S4**). The strongest loci sharing occurred between *iAD2* and *iAging2* (12 CoS), followed by *iAD1* and *iAging3* (7 CoS), consistent with both genome-wide genetic correlation (**Figure 2**) and fine-mapping overlap patterns (3 95% CS and 2 95% CS, respectively) (**Figure S2**). Multi-GWAS colocalization identified proportionally more shared loci (2.5-3 folds increase) than fine-mapping alone. The correlation between variant level fine-mapping posterior inclusion probability (maxPIP) and colocalization probability (maxVCP) was high (r = 0.90) (**Figure S5**), yet only 31% CoS had overlap with fine-mapped 95% CS, demonstrating that multi-trait analysis elevated many lower-confidence signals to 95% threshold through shared evidence across traits, consistent with previously observed power gains^11^. Indeed, 38.8% non-overlapping CoS can be recovered by fine-mapping when we relax the confidence level to 70% CS coverage. Therefore, for downstream analysis we included all 95% CoS as cross-traits effects alongside fine-mapped 95% CS as per trait effects. Hereafter, we refer to these CS and CoS collectively as *loci*.

To assess whether DNE loci disproportionately contribute to disease heritability, we applied stratified linkage disequilibrium score regression (sLDSC) using variants from fine-mapped and colocalized loci as functional annotations, testing whether sparse, mappable signals explain substantial heritability beyond diffuse genome-wide background captured by previous bivariate LDSC analysis. We analyzed six AD GWAS and two aging GWAS (telomere length and mvAge) using variants with maximum variant colocalization probability (maxVCP), an approach demonstrating robust performance^11^ (**Methods**). In AD, we observed significant enrichment for *iAging2* (Enrichment = 97.93, SE = 29.51, p < 0.05), *iAD2* (Enrichment = 49.32, SE = 33.20, p < 0.05), while *iAging5* showed substantial but non-significant enrichment (Enrichment = 42.00, SE = 21.73) (**Figure 2f**). No significant enrichment was detected for aging traits due to the lack of sufficient sample size (number of variants) in 95% CoS genome-wide, although *iAging5*, *iAD1 and iAging3* showed moderate to strong effects (**Figure 2f**). These enrichment patterns are consistent with the bivariate LDSC results, indicating that *iAD2* and *iAging2* jointly contribute to AD pathology, *iAD1* and *iAging3* primarily reflect normal aging processes, and iAging5 appears to influence both. These results establish DNE utility as intermediate phenotypes for gene discovery.

Given DNE loci disproportionately contribute to disease heritability, we leveraged multi-trait colocalization to boost power for identifying endpoint GWAS loci, particularly those shared with DNEs and potentially explained through intermediate imaging phenotypes. We performed multi-GWAS colocalization across 16 GWAS traits, including seven DNE traits and endpoint GWAS for Alzheimer’s disease (AD), aging (telomere length and mvAge), and Parkinson’s disease (PD). For endpoint outcomes, we identified 5,530 AD variants across 101 loci (95% CoS or 95% CS), 4,338 aging variants across 283 loci, and 1,748 PD variants across 16 PD loci. The large variant counts reflect complex LD structure, particularly the *MAPT* locus (chr17:45383525-50162864) capturing 1,561 variants across *iAging3*, AD and PD (**Figure S6**). All 95% CoS reached suggestive significance (p < 1×10⁻⁵), with 64.94%, 87.50%, and 44.62% reaching genome-wide significance (p < 5 ×10⁻⁸) for AD, aging, and PD respectively (**Figure S7**). Among these, 2,224 variants within 13 loci (denoted as orange in **Figure 2**) colocalized across endpoint outcomes and DNEs, representing loci where DNE integration enables finer dissection of endpoint genetic architecture. In comparison, naive overlap of 95% CS between endpoints and DNEs identified only 1 loci. We prioritized these 13 loci as providing strong variant-level evidence linking DNEs to clinical outcomes, which will contribute in a multi-tiered gene prioritization framework described later.

### Brain molecular QTL integration reveals cellular processes underlying neuroimaging endophenotypes

Having established DNE genetic architecture at the variant level, we next characterized the biological processes underlying each DNE before examining their connections to clinical endpoints. We integrated brain molecular QTL data from the FunGen-xQTL consortium, encompassing 48 molecular contexts across bulk and single-nucleus expression, splicing, proteomic, glycoproteomic, metabolomic, and epigenomic QTLs from ROSMAP, MSBB, and Knight ADRC cohorts (n=1,183 total) **(Table S2)**, with DNE GWAS summary statistics through complementary variant-level and gene-level approaches (**Figure 1**; **Methods**). Our xQTL data demonstrated high coverage: more than 60% of DNE loci overlapped with at least one xQTL locus across the 48 contexts (**Figure S8**). We applied pretrained prediction models from FunGen-xQTL to DNE GWAS for transcriptome-wide association studies (TWAS), then integrated TWAS with xQTL colocalization following the putative causal gene (PCG) analysis framework integrating colocalization and TWAS via the *INTACT* method^12^, which requires convergent variant-level and gene-level evidence to reduce false-positives particularly due to linkage disequilibrium (LD) hitchhiking when multiple genes are involved in a locus (**Methods**).

After integrating both lines of evidence, we identified gene sets for each DNE, with the largest for *iAging5* (147 genes), *iAging1* (127 genes), and *iAD2* (97 genes) (**Figure 2**, **Table S8**). High gene-level sharing was observed between *iAD2* and *iAging2* (16 genes), and between *iAD2* and *iAging5* (11 genes), mirroring the loci-level colocalization patterns from the previous section and suggesting these trait pairs capture related neurobiological processes.

We performed Gene Ontology (GO) and KEGG enrichment analysis to annotate DNE (**Methods**). xQTL-derived gene sets provided substantially more specific enrichments than positional mapping of nearby genes (**Figure S9, Table S9–11**), illustrating that functional genomic integration beyond conventional GWAS loci are biologically informative. We focused interpretation on pathways reaching statistical significance (FDR adjusted p < 0.1).

Pathway enrichment defined distinct cellular processes for each DNE (**Figure 3**, **Table S9**). In summary, *iAD1* genes showed enrichment in axolemma, microtubule-associated complex, and tubulin binding, indicating cytoskeletal and axonal transport dysfunction. *iAD2* genes implicated glutamate receptor-mediated excitotoxicity, neuroinflammation, and blood-brain barrier disruption through adherens junction and immune-related pathways. Joint analysis of *iAD1* and *iAD2* strengthened ciliary basal body and cell projection membrane enrichment (**Figure S10, Table S12**). *iAging1* genes were enriched in high-density lipoprotein particle and transport vesicle membrane, suggesting disrupted lipoprotein vesicle transport under ER stress. *iAging2* genes, genetically correlated with *iAD2*, showed enrichment in protein deubiquitination and glutamatergic synapse pathways in joint enrichment, implicating dysregulated ubiquitin-mediated protein degradation at excitatory synapses. *iAging3* genes were enriched in autophagy, VEGF signaling, and hormone secretion, suggesting autophagy impairment and extracellular vesicle dysfunction. Another pair of DNEs with high genetic coherence, *iAD1* and *iAging3*, show shared-gene enrichment in cell projection. This aligns well with the joint enrichment observed for the two AD-derived DNEs in the GO/KEGG analyses and with the AD enrichment detected in the MSigDB Hallmark analysis (**Figures S10–11**). *iAging4* genes showed suggestive evidence in ubiquinone and ketone biosynthetic processes, and significant mitochondrial pathway enrichment when jointly analyzed with *iAD2*, indicating mitochondrial metabolic adaptation during aging complementary to function captured by *iAD2*. *iAging5* genes demonstrated the strongest enrichments among all DNE in peptide antigen binding, ER-to-Golgi transport vesicle membrane, T cell-mediated immunity, and focal adhesion assembly, indicating immune activation, ER-Golgi transport failure, and blood-brain barrier dysfunction potentially through impaired endothelial cell adhesion.

**Figure 3.**
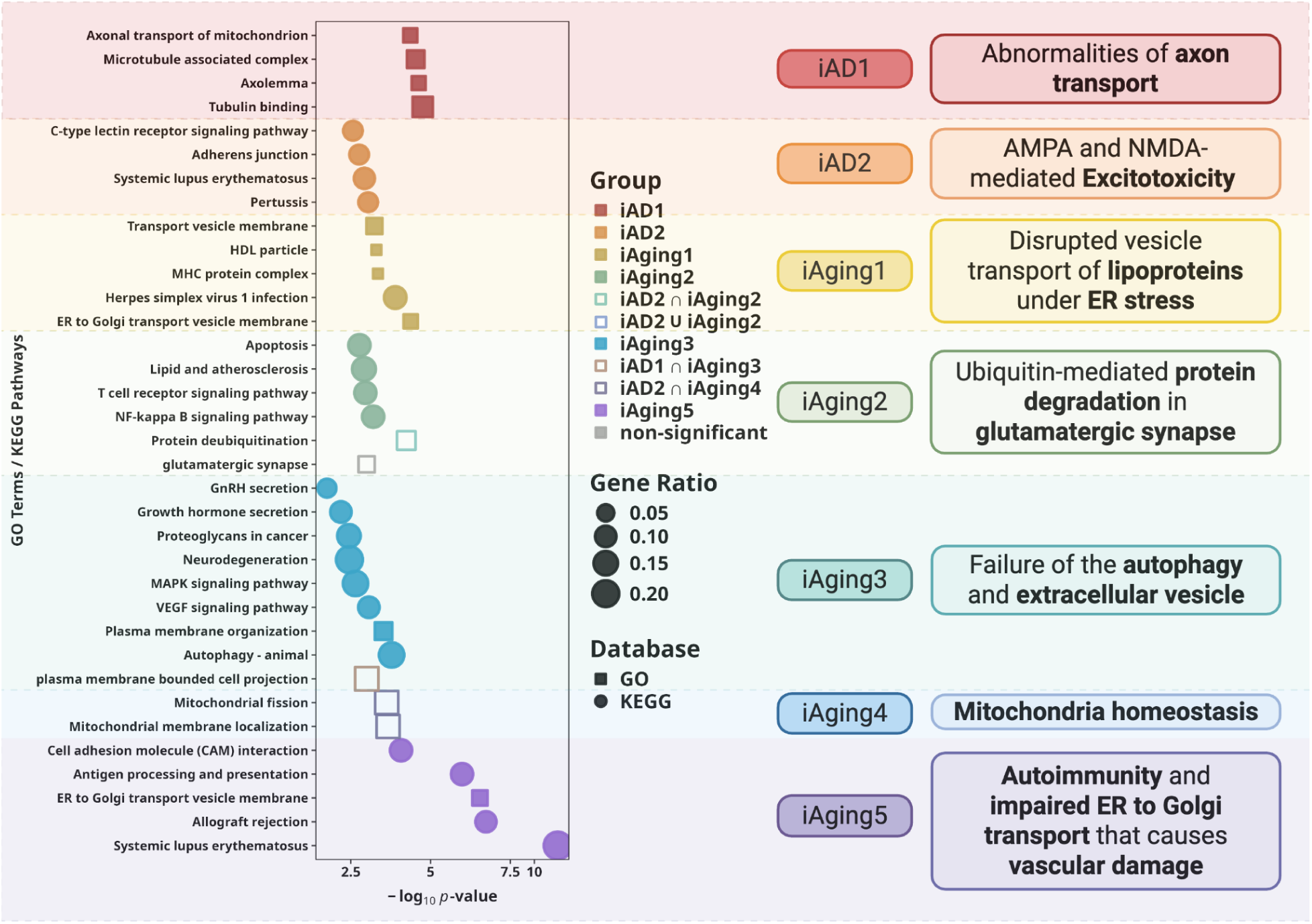
xQTL gene targets dissect distinct cellular processes underlying each DNE. GO and KEGG enrichment of genes from integration of xQTL colocalization and TWAS define biological signatures. Dot size indicates gene ratio; color indicates different DNE with statistical significance (FDR<0.1). Each DNE captures specific cellular responses: *iAD1* represents cytoskeletal and axonal transport dysfunction while *iAD2* reflects glutamate receptor–mediated excitotoxicity; *iAging1* reflects disrupted vesicle transport of lipoproteins under ER stress; *iAging2* indicates dysregulated ubiquitin-mediated protein degradation in excitatory neurons with convergence on glutamatergic synapses; *iAging3* highlights impairments in MAPK signaling, glial differentiation, and autophagy, with extracellular vesicles as potential mediators; *iAging4* points to mitochondrial metabolic adaptation and altered dynamics; and *iAging5* reveals ER-Golgi transport failure and auto-inflammatory states promoting potential vascular damage. Two AD-related DNEs jointly implicate additional pathways involving the ciliary basal body. Collectively, these results show that each DNE represents a distinct but interconnected cellular response underlying aging and AD.

Collectively, xQTL integration biologically characterized each DNE, establishing them as interpretable intermediate phenotypes for mechanistic dissection of disease. With each DNE now defined by specific cellular processes, we leveraged these biological annotations to interpret gene-disease and gene-endpoint connections through DNE-mediated pathways in subsequent analyses, with a particular focus on genes relevant to excitotoxicity and neuroinflammation-mediated vascular damage as these are biological processes annotated to DNE whose genetic correlation are highest with AD (**Figure 2e–f, Figure S11**).

### Multi-tiered evidence framework prioritizes genes linking DNEs to disease endpoints

We next identified which DNE-associated genes influence disease endpoints, by applying a similar variant– and gene-level integration framework to endpoint GWAS combining xQTL colocalization with TWAS across AD, aging (telomere length and multivariate integration of aging traits including healthspan, parental lifespan, exceptional longevity, EAA and frailty), and PD, to prioritize candidate genes where genetic variation influences both brain structure and clinical outcomes. DNEs exhibited substantial gene-level overlap with disease endpoints. The largest overlap was with telomere length (238 shared genes), with *iAging5* contributing most (64 genes), followed by *iAging3* (41 genes), *iAD2* (27 genes), and *iAging1* (17 genes) (**Figure 2g**). Notably, aging-related DNEs (*iAging1-5*), derived from multiple cohorts including UK Biobank capturing population-level brain aging, showed larger overlaps with AD than AD-specific DNEs (*iAD1*: 0 genes; *iAD2*: 17 genes) derived from clinical cohorts to capture disease-specific atrophy. AD showed more modest overlap (51 genes total), with largest contributions from *iAging3* (26 genes), *iAD2* (18 genes), and *iAging5* (9 genes). A three-way intersection of AD, telomere length, and DNEs encompassed 14 genes, suggesting shared genetic architecture between AD and aging dissectable through DNE-mediated pathways.

While integrating DNEs as intermediate phenotypes enhances power, it introduces complexity in systematically weighing multi-scale evidence across molecular contexts, endophenotypes, and endpoints. We therefore developed empirical prioritization rules categorizing candidate genes into evidence-based tiers (**Table 1**, **Figure 1e**). **Tier 1** (27 genes) included genes supported by both loci-level evidence (colocalization) and gene-level evidence (TWAS) in both endpoints and DNEs, representing the strongest convergent evidence. Among Tier 1 genes, 27 mapped to variants within 10loci colocalized between DNEs and endpoints previously identified, including *TNIP1*, *NBEAL1*, and *CBY1*. **Tier 2** (35 genes) comprised genes with both loci-level and gene-level evidence in DNEs plus gene-level evidence in endpoints, including *SEZ6L2* and *NSF*. **Tier 3** (147 genes) included genes with either loci-level or gene-level evidence in DNEs alongside gene-level evidence in endpoints, such as *MAP4* and *DNAJC5*. While Tier 1 represents highest-confidence targets, Tiers 2–3 capture additional genes that, when connected through shared DNE annotations or pathways, construct comprehensive mechanistic hypotheses. Mendelian randomization (MR) was used on endpoints as a screening criterion for tier assignment and also was applied to complement putative causal gene (PCG) analysis in selected gene targets as additional validation, with results presented in **Table S16–17**.

**Table 1.**
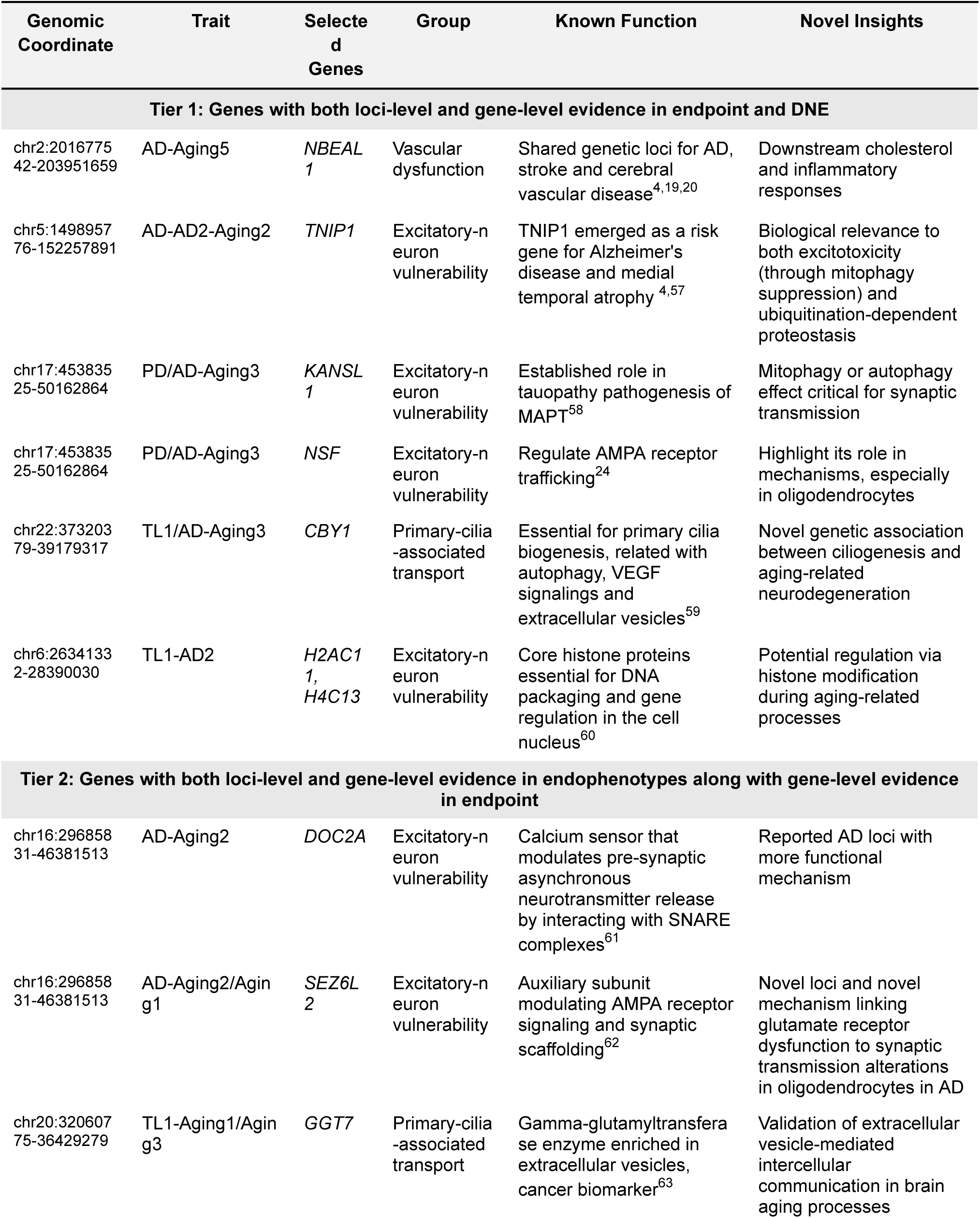
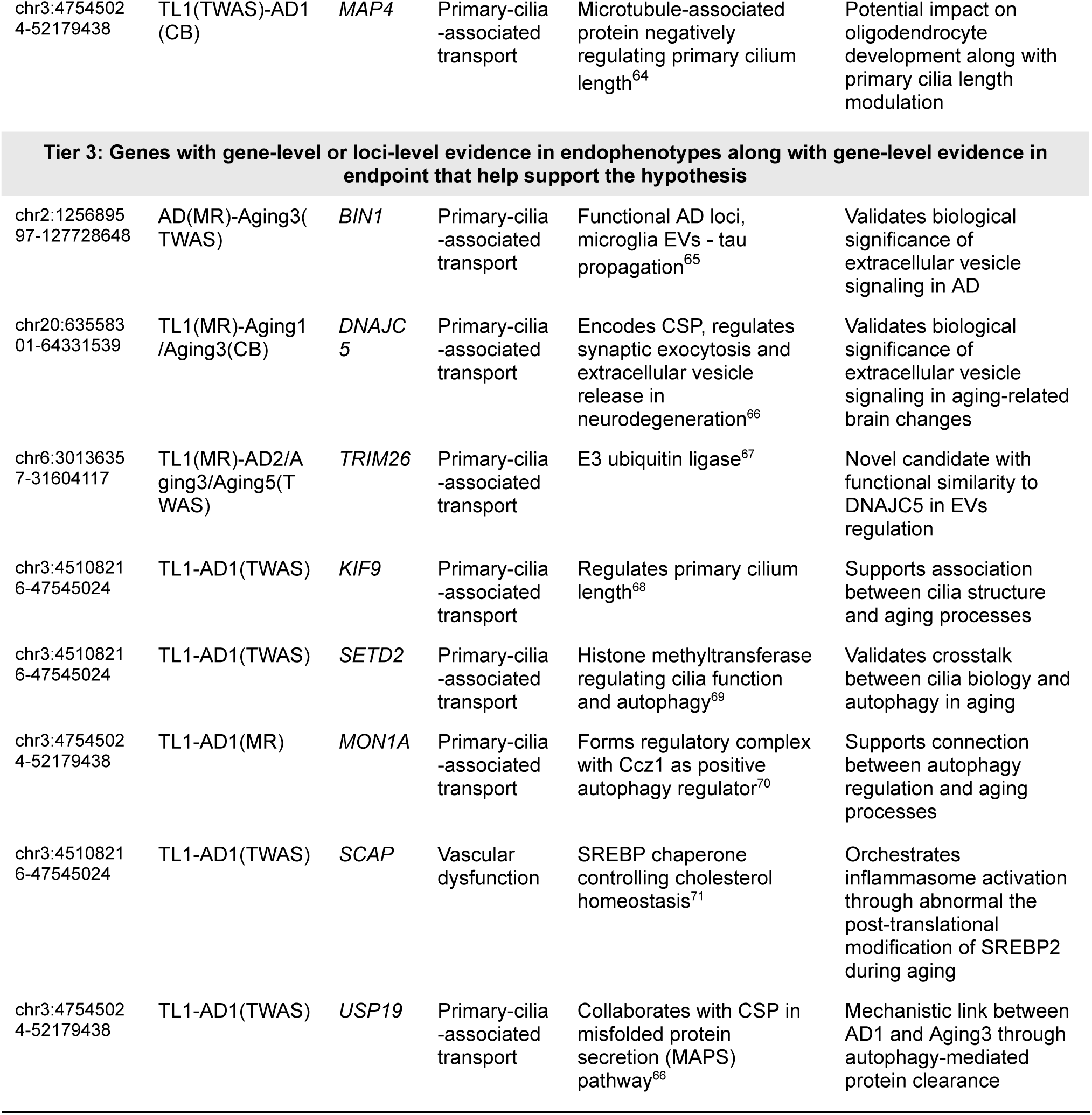
Candidate genes in regulating brain aging with tiers of evidence.

Integrating evidence across tiers and DNE annotations revealed three gene clusters connecting brain endophenotypes to disease outcomes. The synaptic dysfunction cluster encompasses genes mediating mitophagy suppression (*TNIP1*, *KANSL1*; Tier 1), glutamate receptor dysregulation (*NSF*; Tier 1 and *SEZ6L2*; Tier 2), and calcium imbalance (*DOC2A*; Tier 2), linking *iAD2* and *iAging2* to AD through excitotoxicity. The vascular-cholesterol cluster includes *NBEAL1* (Tier 1) and *SCAP* (Tier 3), connecting *iAging5* to AD and vascular dementia through cholesterol dysregulation. The primary cilia cluster comprises *CBY1* (Tier 1) plus *MAP4*, *BIN1*, *DNAJC5*, *TRIM26*, *GGT7*, and four additional genes (Tiers 2–3), linking *iAD1* and *iAging3* to both AD and aging (telomere length) via autophagy-extracellular vesicle pathways. For genomic loci containing multiple candidate genes, we applied cell-type-specific PCG analysis integrating DNE and endpoint evidence to distinguish putative causal genes from LD hitchhikers. Genes meeting stringent PCG criteria with strong mechanistic support are presented in main text, while genes in complex loci (such as the *MAPT* region on chr17) are detailed in **Supplementary Notes** with locus– and context-specific rationale.

In subsequent sections, we present detailed evidence for these gene clusters, focusing on loci with complete xQTL-DNE-endpoint evidence that reveal underexplored regulatory pathways. Established AD genes where our framework provides DNE-mediated annotations are summarized in **Table S12**. We leverage cell-type-resolved xQTL data to dissect locus-specific mechanisms, examining directional effects across cellular contexts, with *in silico* validation using orthogonal data (AD pathology, cross-species models) to generate mechanistic hypotheses connecting pre-symptomatic brain alterations to disease.

### Cell-specific roles of synaptic dysfunction and excitotoxicity in neurodegenerative disorders

#### Mitophagy suppression in excitatory neurons

Cell-type-resolved DNE-xQTL integration identified *TNIP1* (chr5, Tier1) with excitatory neuron-specific contribution to excitotoxicity connecting *iAD2* and *iAging2* to AD. *TNIP1* encodes a regulator involved in mitophagy and NF-κB mediated inflammatory control, with known roles in cellular stress and mitochondrial quality regulation (**Table 1**). Specifically, *TNIP1* expression is positively associated with AD (p_TWAS_ = 3.2 × 10^−11^), *iAD2* (p_TWAS_ = 5.1 × 10^−8^), and *iAging2* (p_TWAS_ = 8.1 × 10^−7^) in excitatory neurons (**Figure 4a**), and the lead variant rs871269 (chr5:151052827:C>T) colocalized across these traits and two excitatory neuron eQTL datasets (**Figure 4b**). Consistent with its association with lower AD risk and DNE scores, rs871269 also demonstrated a negative association with TDP-43 burden in ROSMAP samples (nominal p = 0.02) (**Figure 4c**). Complete multi-omic evidence, variant-level colocalization with multiple AD GWAS datasets are provided in **Supplementary Note S4.**

**Figure 4.**
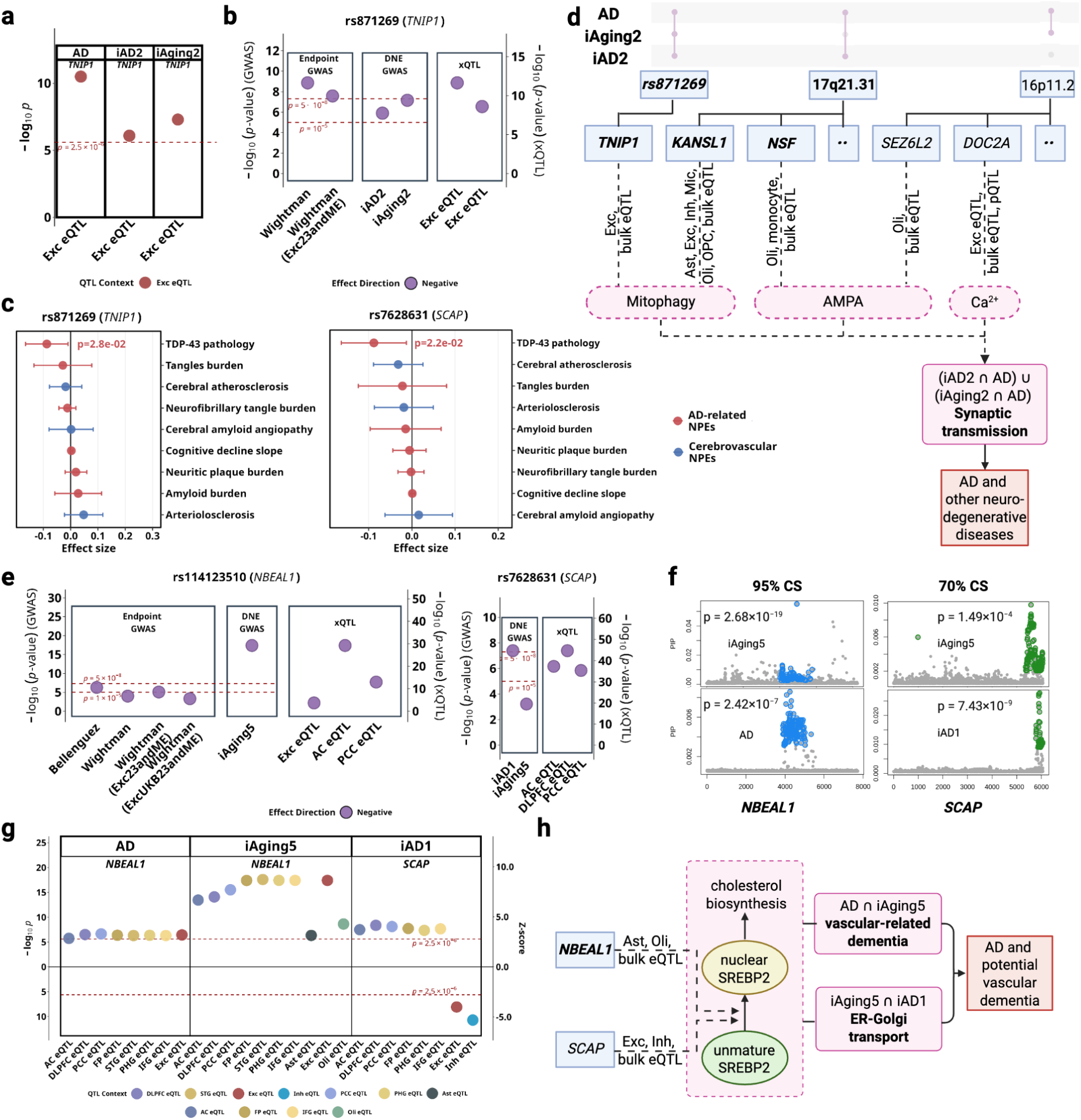
*iAD1* and *iAD2* implicate excitotoxicity and cholesterol dysregulation in AD pathogenesis. **a-d** shows the synaptic transmission regulatory hub; **e-h** shows the cholesterol/vascular regulatory hub. **(a)** TWAS p-values for *TNIP1* across significant tissues/cell types denoted with different colors. (positive associations in excitatory neurons) **(b)** Lead *TNIP1* variant (rs871269) colocalization across datasets with AD. **(c)** Pathology associations for lead variants for *TNIP1* and *SCAP*. **(d)** Possible mechanistic model involving key biological processes in synaptic transmission: mitophagy, glutamate receptor signaling, and calcium imbalance. **(e)** Lead *NBEAL1* variant (rs114123510) colocalization across AD and *SCAP* variant (rs7268631) across *iAD1*, both involving *iAging5*, with (**f**) fine-mapping 95% and 70% credible sets shown for these variants, respectively. **(g)** TWAS p-values and Z-scores for *NBEAL1* and *SCAP* across significant tissues/cell types. **(h)** Possible mechanistic model involving modulation of SREBP maturation to influence AD and other vascular-related dementia.

#### AMPA receptor dysregulation and mitophagy at the MAPT locus

The 17q21.31 region harbors 22 Tiers 1–3 genes in extensive LD due to a large inversion polymorphism^13^, making conventional gene prioritization challenging. Previous GWAS suggest pleiotropic effects at this locus across neurodegenerative disorders including AD, PD, and amyotrophic lateral sclerosis (ALS)^7,14,15^. Cell-type-specific putative causal gene (PCG) analysis (**Methods**) integrating DNE and AD evidence identified *KANSL1*, *NSF*, and *CRHR1* as candidate causal genes at this locus, with distinct cell-type contributions (**Supplementary Note S1**). *KANSL1*, part of the NSL chromatin-modifying complex linked to mitochondrial and mitophagy regulation (**Table 1**), showed protective associations with AD and *iAD2* across excitatory neurons and oligodendrocytes. *NSF*, an ATPase required for AMPA receptor trafficking (**Table 1**), displayed oligodendrocyte-specific associations with DNE traits and exhibited opposing directions between *iAD2* and symptomatic AD. *CRHR1*, which modulates glutamatergic and stress-responsive signaling pathways (**Table 1**), showed significant associations in oligodendrocytes and excitatory neurons across DNE and AD endpoints. These genes collectively implicate both neuronal and glial mitochondrial dysfunction and glutamate receptor dysregulation as converging mechanisms at this genomically complex locus.

#### Oligodendrocyte AMPA receptor scaffolding at chromosome 16

The 16p11.2 region contains 26 Tiers 2–3 genes in high linkage disequilibrium, presenting similar prioritization challenges (**Table S13**). Cell-type-specific PCG analysis identified *SEZ6L2*, *DOC2A*, and *MAPK3* with oligodendrocyte-specific associations linking them to *iAging2* (**Supplementary Note S2**). *SEZ6L2* functions as an AMPA receptor scaffolding protein (**Table 1**) and showed the strongest oligodendrocyte-specific evidence, with opposing directional patterns between *iAging2* and AD that parallel those observed for *NSF*. *DOC2A*, which encodes a calcium sensor modulating glutamate release (**Table 1**), showed complex context-dependent associations across mRNA and protein levels in AC, DLPFC and excitatory neurons. *MAPK3*, relevant to p38-MAPK signaling, showed pleiotropic effects across AD, telomere length, *iAD2*, *iAging2* in oligodendrocytes and microglia (**Figure S12, Table S14**). Together, these genes reveal potential oligodendrocyte contributions to AMPA receptor-mediated excitotoxicity through receptor scaffolding, calcium sensing, and stress signaling mechanisms, expanding traditional neuron-centric models of excitotoxicity in AD.

Collectively, our findings suggest mitochondrial dysfunction and glutamate receptor dysregulation may synergistically contribute to excitotoxicity in neurons and oligodendrocytes, and are potentially relevant to pre-symptomatic changes to AD (**Figure 4d**).

### Cell-type-resolved xQTL suggests NBEAL1-mediated vascular-cholesterol dysregulation linking *iAging5* and *iAD1* to AD

Cerebrovascular pathology co-occurs in over 80% of AD cases^16^, but the cell-type-specific molecular mechanisms remain poorly characterized. Although our xQTL datasets lack cerebral endothelial cells, the biological annotations of *iAging5* (ER-Golgi transport failure, BBB disruption, vascular inflammation) and *iAD1* (axonal transport dysfunction) suggested vascular mechanisms warranting investigation. Our cell-type-resolved xQTL integration identified *NBEAL1* (Tier 1) and *SCAP* (Tier 3), which have been reported to implicate SREBP2-mediated cholesterol homeostasis as a mechanism connecting DNEs to cerebrovascular contributions to AD pathogenesis^17,18^.

*NBEAL1* is a regulator of vesicular trafficking and endolysosomal function (**Table 1**), exhibits positive associations between gene expression and AD across six brain regions (p_TWAS_ < 2.5 × 10⁻^6^ in AC, DLPFC, PCC, FP, STG, IFG) with stronger associations for *iAging5* in the same regions (**Figure 4e**; also **Table S14** for TWAS p-values and **Table S15** for brain region abbreviations). Cell-type-resolved analysis demonstrated positive associations in excitatory neurons (AD: p_TWAS_ = 3.60 × 10⁻⁷; *iAging5*: p_TWAS_ = 3.87 × 10⁻¹⁸), astrocytes (*iAging5*: p_TWAS_ = 1.3 × 10⁻⁶), and oligodendrocytes (*iAging5*: p_TWAS_ = 2.3 × 10⁻⁹). GTEx data demonstrate abundant *NBEAL1* expression in arterial tissue despite the absence of cerebral endothelial cells in our xQTL datasets (**Figure S13**). The lead variant rs114123510 (chr2:202966489:T>A) colocalized across *iAging5*, AD and brain eQTL contexts including bulk tissues (AC, PCC) **and excitatory neurons** (**Figure 4f**).

*SCAP*, known to escort SREBPs from the ER to the Golgi (**Table 1**), exhibits positive associations between gene expression and *iAD1* in bulk brain tissue (p_TWAS_ < 2.5 × 10⁻^6^ in DLPFC, PCC, FP, PHG, IFG), but negative associations in excitatory neurons (p_TWAS_ = 2.2 × 10⁻⁸) and inhibitory neurons (p_TWAS_ = 5.3 × 10⁻⁸) (**Figure 4e**). The lead variant rs7628631 (chr3:46,921,725-47,513,711) colocalized across *iAD1*, *iAging5*, and brain eQTL datasets (AC, DLPFC and PCC, **Figure 4f**). Despite weak marginal association with *iAging5*, fine-mapping nonetheless placed these variants in a 70% credible set (**Figure 4g**).

*NBEAL1* and *SCAP* encode functionally coupled proteins regulating SREBP2 processing with established roles in endothelial cholesterol homeostasis and vascular disease (**Table 1**). Our integration reveals that both genes connect to AD through DNE capturing vascular dysfunction: *NBEAL1* associates primarily with *iAging5*, annotated for ER-Golgi transport failure, BBB disruption, and vascular inflammation (**Figure 3**); *SCAP* shows associations with both *iAD1* (axonal transport dysfunction) and *iAging5* (vascular dysfunction). The convergent evidence linking both genes to the SREBP2 pathway, combined with *NBEAL1* proximity to cerebral small vessel disease loci^19,20^, implicates cerebrovascular cholesterol dysregulation as a mechanism contributing to AD pathogenesis (**Figure 4h**). Additional genes at the *NBEAL1* locus (*CARF*, *ICA1L*, *WDR12*) showed variant-level colocalization at the *NBEAL1* locus but lacked consistent gene-level support from PCG analysis and clear mechanistic connection to vascular dysfunction or cholesterol pathways; these results are detailed in **Supplementary Note S3**.

### Primary cilia integrate autophagy and extracellular vesicle pathways linking DNE to telomere length and AD

Primary cilia are sensory organelles that regulate cellular homeostasis through intraflagellar transport and signaling pathways, with established roles in aging. DNE-xQTL integration identified ten genes spanning 7 genomic loci associated with telomere length and/or AD that converge on ciliary function through two possible mechanisms: extracellular vesicle-mediated intercellular communication (*BIN1*, *CBY1*, *DNAJC5*, *TRIM26*, *GGT7* via *iAging3*), and structural regulation of cilia and autophagy (*MAP4*, *KIF9*, *SETD2*, *MON1A*, *USP19* via *iAD1*) (**Figure 5g**). LD structure between lead variants from colocalized sets of genes (**Table S13**) and prioritization rationale are detailed in **Supplementary Note S4**.

**Figure 5.**
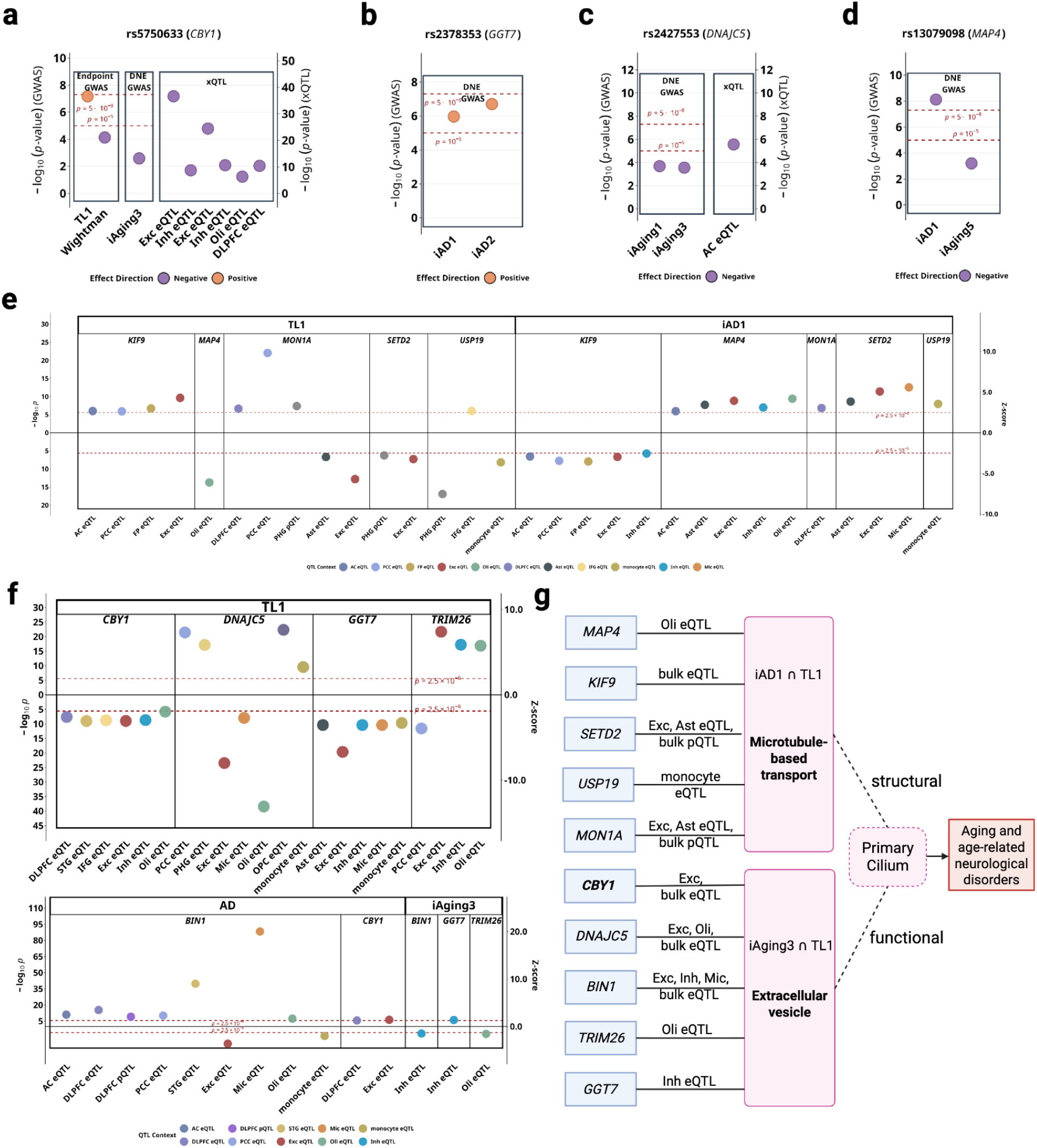
*iAD1* and *iAging3* implicate primary cilia dysfunction in aging and AD pathology. **(a-d)** Lead variants colocalization across endpoint and DNE GWAS, and brain xQTL datasets for *CBY1*, *GGT7*, *DNAJC5* and *MAP4*. **(e)** TWAS p-values and Z-scores for *iAD1*-telomere length genes (*MAP4*, *KIF9*, *SETD2*, *MON1A*, *USP19*) across significant tissues/cell types denoted by different colors. **(f)** TWAS p-values and Z-scores for *iAging3*-telomere length genes (*CBY1*, *DNAJC5*, *BIN1*, *TRIM26*, *GGT7*) across significant tissues/cell types. **(g)** Possible mechanistic model involving 10 telomere length and/or AD-associated genes linked to *iAD1* or *iAging3*, all structurally or functionally related to primary cilia.

#### Ciliogenesis and extracellular vesicle communication

*CBY1* (Tier 1), an essential gene for primary ciliogenesis (**Table 1**), showed positive associations with telomere length and negative associations with AD in excitatory neurons (**Figure 5e**). The lead variant rs5750633 (chr22:38597462:A>G) colocalized across telomere length, AD, *iAging3*, and brain eQTL datasets including DLPFC, excitatory neurons, inhibitory neurons, and oligodendrocytes (**Figure 5a**), with multi-trait colocalization supported by strong molecular QTL evidence. Despite weaker marginal GWAS *iAging3* association, we consider this DNE relevant because its annotated biological processes involve autophagy impairment and VEGF-related processes (**Figure 3**, **Table 1**). *BIN1*, a tau-interacting protein involved in extracellular vesicle-mediated tau trafficking and the second most strongly associated AD locus after *APOE* (**Table 1**), demonstrated negative associations with *iAging3* in inhibitory neurons and with AD in excitatory neurons, but positive associations with AD in other contexts including microglia, revealing cell-type-dependent roles in EV-mediated tau propagation. Additional genes (*DNAJC5*, *TRIM26*, *GGT7*) connect telomere length to neurodegeneration through EV-mediated pathways, extending known mechanisms of tau spreading and EV regulation to aging processes through *iAging3* (**Table 1**). Complete TWAS statistics, variant-level colocalization, and functional details for all EV communication genes are provided in **Supplementary Note S3**.

#### Structural ciliary regulation and autophagy

*MAP4*, known to regulate primary cilium length through modulation of axonemal microtubule stability (**Table 1**), demonstrated cell-type-specific associations exclusively in oligodendrocytes: negative with telomere length and positive with *iAD1* (**Figure 5e**). *MON1A* and *USP19*, two autophagy-related genes involved in autophagosome maturation and protein quality control, showed distinct associations with telomere length and *iAD1* where *MON1A* showed positive associations with both telomere length and *iAD1* in DLPFC, while *USP19* showed negative associations with telomere length and positive associations with *iAD1* in monocytes (**Figure 5a**). Additional supporting genes (*KIF9*, *SETD2*), along with complete statistical evidence and variant-level colocalization for all structural regulation genes, are detailed in **Supplementary Note S3**.

## DISCUSSION

Our study demonstrates that integrating imaging-derived brain endophenotypes with multi-omics molecular QTL profiles and disease GWAS provides a coherent genetic framework for interpreting early neurobiological changes. By linking imaging-derived endophenotypes to gene regulation across multiple molecular modalities, the DNE-xQTL framework identifies mechanisms underlying pre-symptomatic brain alterations and their contributions to neurological disease. Our *DNE-xQTL framework* addresses critical gaps in current approaches for understanding disease mechanisms. GWAS identify disease risk loci without revealing how variants influence brain changes, and xQTL studies link genetics to molecular phenotypes without capturing *in vivo* neurobiology. However, our integration simultaneously characterizes the molecular architecture of pre-symptomatic brain alterations and connects them to disease through convergent genetic evidence.

Specifically, using DNEs as intermediate phenotypes boosts statistical power to detect disease-relevant genes, especially at loci with modest AD signals (P < 1×10⁻⁵) that gain support through shared imaging and molecular evidence. Secondly, integrating convergent genetic and functional evidence at variant– and gene-level through systematic statistical framework enables gene prioritization at complex loci to support focused mechanistic hypotheses. Third, cell-type-resolved xQTL integration revealed context-specific mechanisms, demonstrating that the same gene may exhibit divergent effects across cell types or disease stages, providing insights into disease etiology. Fourth, our framework can identify fine-mapped loci across molecular traits, endophenotypes and endpoints, prioritizing from thousands of associated variants a handful candidates as instrumental variables for causal inference and mechanistic studies.

Together, these advances suggests three putative regulatory hubs connecting pre-symptomatic brain changes to neurodegeneration: a synaptic dysfunction hub implicating oligodendrocyte contributions to glutamate receptor-mediated excitotoxicity; a vascular-cholesterol hub linking BBB dysfunction to cerebrovascular dementia through SREBP2-mediated cholesterol dysregulation; and a primary cilia hub positioning cilia as integrators of cellular aging and intercellular protein propagation.

### SREBP2-mediated cholesterol homeostasis

The proposed vascular-cholesterol hub demonstrates how SREBP2-mediated cholesterol homeostasis connects neuroimaging patterns potentially relevant to blood-brain barrier dysfunction and cerebrovascular inflammation. SREBP2 acts as the master regulator of cholesterol biosynthesis and also involved in immune responses and neurodegeneration progression. Within the vascular-cholesterol hub, convergent genetic evidence points to a regulatory locus on chromosome 2 containing *NBEAL1*, *CARF*, *ICA1L*, and *WDR12*, among which *NBEAL1* was prioritized based on convergent lines of evidence: strongest TWAS associations across six brain regions and three cell types (excitatory neurons, astrocytes, oligodendrocytes); genomic proximity to GWAS loci for lacunar stroke and cerebral small vessel disease^19,20^; and involvement in endothelial cholesterol homeostasis (e.g. upregulated heart endothelial expression in obesity^21^). Follow up *in silico* analysis in ROSMAP and GTEx further strengthened these roles. *NBEAL1* enhances SREBP2 proteolysis in endothelial cells, promoting cholesterol biogenesis in HUVECs^17^. Notably, the observed associations with *iAging5* in astrocytes and oligodendrocytes are particularly relevant given that these cell types are the primary sources of brain cholesterol^22^. Under physiological conditions, *SCAP* traffics SREBP2 for proteolytic activation and nuclear signaling that maintains cholesterol homeostasis^22^, and also transports NLRP3 for controlled inflammasome activation^18^. We hypothesize that pathologically enhanced *NBEAL1*-mediated SREBP2 proteolysis drives excessive SCAP-SREBP2 activity concurrently with NLRP3-dependent inflammation, ultimately contributing to neurodegenerative outcomes. This cascade may result in hypercholesterolemia and chronic neuroinflammation that compromises vascular integrity of small cerebral vessels, ultimately promoting cognitive decline characteristic of vascular dementia (**Figure 6b**). The cell-type-resolved convergence of genetic evidence from *iAging5* (annotated for ER-Golgi transport failure, BBB disruption, and vascular inflammation) and *iAD1* (microtubule dysfunction), together with vascular disease AD and aging GWAS signals, alongside endothelial cholesterol biology, generates a testable hypothesis with candidate genetic variants as tools for functional validation. Understanding these pathways may deepen our knowledge on vascular dementia, with clinical translation targeting cholesterol dysregulation in early disease stages, with *iAD1* and *iAging5* serving as a screening endophenotype to identify patients at risk for this vascular mechanism.

**Figure 6.**
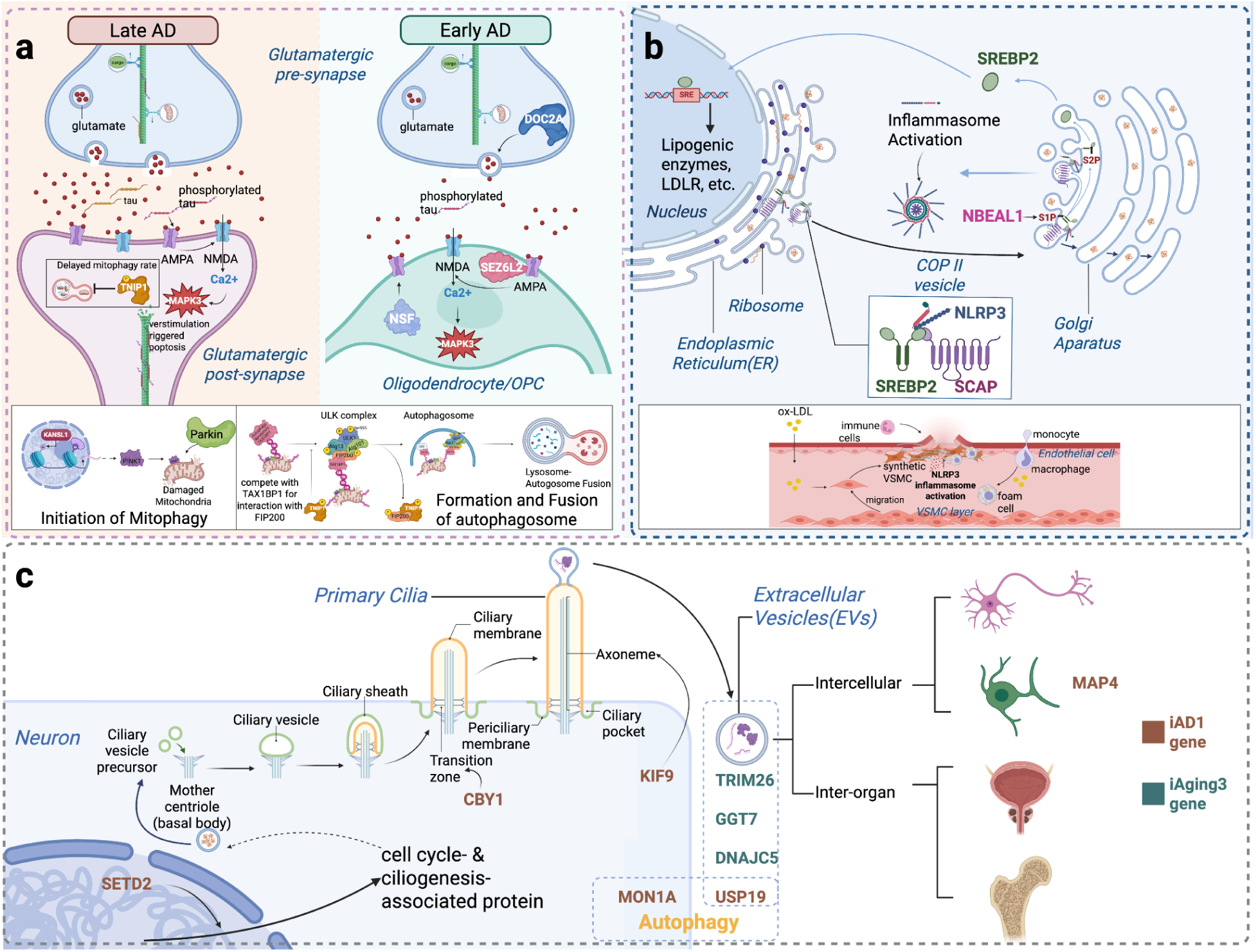
Summary of hypotheses for DNE-mediated pathways in AD and aging. **(a)** Oligodendrocytes in excitotoxicity: Hippocampal synaptic transmission declines in aging through reduced ubiquitin-mediated protein regulation (*iAging2*)^51^. In AD, glutamate excitotoxicity via AMPA/NMDA receptors (*iAD2*) drives synaptic dysfunction^52^. *DOC2A* promotes synaptic transmission by modulating calcium level, showing associations in both *iAging2* and AD. *KANSL1* modulates AMPA receptor breakdown, while *NSF* regulates AMPA receptor trafficking, and *SEZ6L2* serves as AMPA receptor regulators—all showing associations with *iAD2/iAging2* and AD. Mitochondrial dysfunction exacerbates both aging and AD pathways. *TNIP1* suppresses mitophagy, showing positive associations with *iAD2*, *iAging2*, and AD. Critically, *NSF* and *SEZ6L2* expression in oligodendrocytes reveals essential glial contributions to excitotoxicity regulation, expanding beyond neuron-centric models. **(b) Cholesterol dysregulation in vascular dementia:** Under physiological conditions, *SCAP* escorts SREBP2 from ER to Golgi, where S1P and S2P proteases cleave SREBP2 to release its active fragment, which translocates to the nucleus to induce LDL receptor expression and maintain cholesterol homeostasis^53^. *SCAP* also transports NLRP3 for controlled inflammasome activation^54^. In pathological states, *NBEAL1* may enhance SREBP2 proteolysis, potentially driving excessive SCAP-SREBP2 activity that causes aberrant LDL receptor upregulation and cholesterol accumulation. This could trigger NLRP3-dependent inflammation, resulting in hypercholesterolemia and chronic neuroinflammation that may compromise vascular integrity of small cerebral vessels, ultimately promoting cognitive decline and vascular dementia. **(c) Primary cilia as aging regulators:** Ten telomere length-associated genes converge on ciliary function through two mechanisms. *MAP4*, *KIF9*, *SETD2*, *MON1A*, and *USP19* participate in intraflagellar transport, which is analogous to axonal transport, and may regulate cilia-autophagy interactions^55^, with *MAP4* exclusively expressed in oligodendrocytes, suggesting potential roles in cilia dynamics during oligodendrocyte maturation^56^. *CBY1*, *DNAJC5*, *BIN1*, *TRIM26*, and *GGT7* mediate extracellular vesicle formation and secretion for intercellular communication. *CBY1* is essential for ciliogenesis; *DNAJC5* and *USP19* regulate EV-mediated misfolded protein release; *TRIM26* functions in oligodendrocyte EVs, while *BIN1* operates in inhibitory neuron EVs; *GGT7* serves as an EV cargo protein. Collectively, these genes suggest primary cilia may function as central regulators integrating intracellular transport with intercellular communication in aging and neurodegeneration.

### Glutamate receptor dysregulation and mitochondrial dysfunction in excitotoxicity

The proposed synaptic dysfunction regulatory hub implicates glutamate receptor dysregulation and mitochondrial dysfunction in excitotoxicity, which is a fundamental mechanism that initiate and propagate neurodegenerative cascades, with further evidence pointing to oligodendrocyte contributions expanding beyond neuron-centric models. Through DNE-xQTL integration, we prioritized genes within two genomically complex regions where conventional nearest-gene approaches cannot readily distinguish among multiple candidates. At MAPT locus (17q21.31) with extensive LD, we applied PGC analysis (**Methods**) integrating evidence across both DNE traits and disease endpoints, as xQTL and endpoint alone cannot resolve gene prioritization in such complex loci where 54 genes show xQTL-AD association evidence in our tiered genes. This DNE-mediated dissection identified *KANSL1*, *NSF*, and *CRHR1*, with *KANSL1* showing consistent effects across *iAD2*, *iAging2*, and AD, while others showing opposing directional patterns are examined in **Supplementary Notes S1**. These observations suggest convergence of mitochondrial vulnerability and glutamatergic dysregulation that may influence both early excitotoxicity and later disease progression. Another LD-dense region on chromosome 16 with a distinct but related regulatory architecture across 32 genes, shows xQTL-AD association evidence. DNE integration prioritized *DOC2A*, *SEZ6L2*, and *MAPK3* based on oligodendrocyte-specific associations with *iAD2* and *iAging2*. *MAPK3* showed broad pleiotropic effects across multiple contexts, whereas *SEZ6L2* and *DOC2A* displayed directions consistent with compensatory responses in early aging and potential post-transcriptional modulation of calcium homeostasis in glutamatergic synapse dysfunction. Notably, *NSF* and *SEZ6L2, which are* both AMPA receptor regulators, showed oligodendrocyte-specific associations, suggesting that oligodendrocytes may modulate glutamate receptor trafficking and participate in excitotoxic processes traditionally attributed to neurons. This complements traditional neuron-centric excitotoxicity models and suggests glial-targeted therapeutic strategies. *TNIP1* and *KANSL1* further implicate mitochondrial dysfunction through mitophagy suppression. *TNIP1* shows excitatory neuron specific associations that may increase early AD vulnerability through mitophagy suppression, consistent with the mitochondrial pathway enrichment observed for *iAD2* and *iAging4*; while *KANSL1* additionally shows strong PD associations, indicating that mitochondrial dysfunction may represent a shared mechanism across neurodegenerative disorders. These findings reveal possible *MAPT*-independent therapeutic targets through synaptic transmission functions (**Figure 6a**), demonstrating how intermediate phenotypes may enable mechanistic gene prioritization in genomically complex regions. Additional mechanistic interpretation, including epigenetic regulation through chromatin accessibility and opposing bulk versus single-cell effects, is provided in **Supplementary Notes S3 and S5**.

### Intracellular transport and intercellular communication through primary cilia in aging

The proposed primary cilia hub positions cilia as regulators integrating intracellular transport with intercellular communication in aging and neurodegeneration. Primary cilia dysfunction has also been linked to numerous neurological disorders, underscoring their sensitivity to aging and involvement in neurodegenerative processes^23^. Structurally, ciliary axoneme, built from nine microtubule doublets, supports bidirectional intraflagellar transport (IFT) and may contribute to axon transport relevant processes captured by *iAD1*. Functionally, primary cilium is both a broadcaster and a receiver in a complex cellular communication network mediated by EVs, which is implicated by *iAging3*. A total of 145 genes were implicated by DNE-xQTL integration, including intraflagellar transport genes (analogous to axonal transport captured by *iAD1*) and EV pathway genes (implicated in tau propagation captured by *iAging3*), suggest cilia coordinate both cellular homeostasis and intercellular protein propagation. The convergence of *CBY1* (essential for ciliogenesis) and *MAP4* (which increases during oligodendrocyte differentiation as cilia disassemble) situates this genetic architecture within a broader framework connecting ciliary dynamics to axonal transport and myelination biology in aging and AD. In addition, *MON1A* participates in autophagosome maturation and endolysosomal trafficking, whereas *USP19* contributes to *DNAJC5*-dependent EV export of misfolded proteins, illustrating how autophagy-related regulators could interface with the primary cilia hub. This combined statistical and functional evidence generates the hypothesis that primary cilia dysfunction may represent a nexus linking cellular aging to pathological protein propagation in neurodegeneration, suggesting that cilia function not only as sensory organelles but also as integrators of proteostasis and intercellular signaling **(Figure 6c)**. Detailed interpretation of individual genes, their tissue-specific effects, and the autophagy-EV mechanistic links is detailed in **Supplementary Notes S3 and S4**.

### Opposing directional effects in oligodendrocytes

The opposing directional effects observed for several genes (*NSF*, *SEZ6L2*, *DOC2A*, *MAPK3*), *i.e.*, elevated expression with pre-symptomatic traits (*iAD2*, *iAging2*) but reduced expression in symptomatic AD, may reflect stage-dependent regulatory mechanisms. All four genes show their strongest associations in oligodendrocytes, which consistently exhibit early upregulation consistent with their roles in axonal support and buffering excitatory stress. For example, elevated oligodendrocyte-specific expression of *SEZ6L2*, *DOC2A*, *MAPK3* in *iAging2* may represent compensatory responses supporting myelination and synaptic communication under early aging-related stress, in contrast to their reduced expression in symptomatic AD. Similarly, at the 17q21.31 locus, *NSF* showed positive associations with *iAD2* but negative associations with AD. Since *NSF* promotes AMPA receptor surface expression while tau protein inhibits its activity^24^, elevated expression in early stages may increase receptor-mediated excitotoxic vulnerability before tau pathology suppresses its function. These bidirectional patterns raise the possibility that DNE traits reflect transitional or compensatory states distinct from the end-stage pathology observed in symptomatic disease.

This study has several limitations that highlight future directions. As an *in silico* gene prioritization framework leveraging multi-scale genetic data, functional validation through experimental models remains essential to test our mechanistic hypotheses. Our xQTL contexts lack certain disease-relevant cell types, particularly cerebral endothelial cells needed to strengthen vascular mechanisms, and are limited to post-mortem brain tissue that may not capture dynamic regulatory states antemortem. In addition, establishing causality across the complete xQTL-DNE-endpoint pathway remains challenging. While Mendelian randomization has been applied to pairs of traits (DNE-endpoint and xQTL-endpoint), extending causal inference across all three studies sequentially with limited instrumental variables poses methodological challenges for future statistical genetics methods development. Beyond these limitations, this framework opens several promising directions. First, incorporating multi-organ neuroimaging endophenotypes and xQTL data would enable systematic investigation of systemic aging mechanisms beyond the brain. Second, multi-ancestry xQTL studies would enable higher-resolution fine-mapping in complex loci where European-ancestry LD patterns limit variant resolution. Third, establishing longitudinal neuroimaging-molecular cohorts that track individuals from pre-symptomatic to symptomatic stages could directly test stage-dependent mechanisms. In the longer term, linking DNE-derived early brain signatures with molecular pathways may support earlier risk stratification, refine target selection, and inform clinical strategies aimed at intervening before irreversible neurodegeneration occurs.

In conclusion, this gene prioritization and hypothesis-generating framework demonstrates that biologically characterized neuroimaging endophenotypes, when integrated with comprehensive multi-context molecular QTL data, can reveal candidate molecular targets for early intervention in neurodegeneration. By transforming abstract imaging patterns into testable cellular hypotheses with molecular and cell-type resolution, this approach enables biologically motivated gene prioritization in genomically complex regions where conventional GWAS approaches fail. The three regulatory hubs identified provide candidates for experimental validation and therapeutic development targeting pre-symptomatic brain changes before irreversible neurodegeneration occurs.

## METHODS

### Data description

Summary statistics for Dimensional Neuroimaging Endophenotypes (DNEs) and Alzheimer’s disease (AD) from multiple cohorts were obtained from publicly available repositories as described in the original publications. Details of data sources, cohort composition, and sample sizes are provided in **Table S1**. Briefly, DNEs were defined using machine learning models trained on multimodal MRI features to capture disease-related neurobiological dimensions. These imaging-derived traits were developed through a standardized framework and validated across multiple brain disorders, with the trained models applied to UKBB participants (n = 32,829-33,541) to generate continuous DNE scores and GWAS performed adjusting for age, sex, their interactions with age and age-squared, total intracranial volume (ICV), and genetic principal components, yielding associations with common genetic variants. All genomic coordinates were harmonized to the GRCh38 (hg38) reference assembly using LiftOver^25^.

This work is based on summary statistics from multiple molecular quantitative trait locus (xQTL) datasets, including bulk and single-nucleus expression, splicing, proteomic, glycoproteomic, metabolomic, and epigenomic (DNA methylation, histone modification, and single-nucleus Assay for Transposase-Accessible Chromatin) QTLs, were generated by the Functional Genomics Consortium xQTL Working Group (FunGen-xQTL) using data from the Religious Orders Study and Memory and Aging Project (ROSMAP), Mount Sinai Brain Bank (MSBB), and Knight Alzheimer Disease Research Center (ADRC). The combined multiple brain regions include anterior cingulate cortex (AC), dorsolateral prefrontal cortex (DLPFC), and posterior cingulate cortex (PCC) from ROSMAP; and frontal pole (FP), inferior frontal gyrus (IFG), superior temporal gyrus (STG), and parahippocampal gyrus (PHG) from MSBB; and primarily parietal cortex from Knight ADRC as summarized in **Table S2** with details available in the corresponding publications^26–32^.

### Fine-mapping

For DNE and AD GWAS summary statistics described in **Table S1**, we performed the following quality control procedures prior to fine-mapping analysis. Variants with missing alleles, strand mismatches, or duplicates were excluded, and allele orientation was aligned to the linkage disequilibrium (LD) reference panel. The LD reference was estimated from 17,000 unrelated whole-genome-sequenced European individuals from the Alzheimer’s Disease Sequencing Project (ADSP), provided by the Genome Center for Alzheimer’s Disease (GCAD)^33^. Strand-ambiguous SNPs (A/T or C/G), variants with missingness greater than 1%, minor allele frequency (MAF) below 0.0025, or not shared between GWAS and genotype-based LD reference panels were removed. Additional quality control was performed to detect and remove variants with inconsistent z-scores or abnormal LD patterns. For each LD block, z-scores were jointly modeled with the local LD matrix under the SuSiE^34^ framework to identify discrepancies between the statistical signal (z) and the corresponding LD structure (R). Variants exhibiting outlier behavior were excluded from both the summary statistics and LD matrix to ensure internal consistency. Missing or filtered variants were then imputed using the RAISS^35^ algorithm, which reconstructs missing z-scores by leveraging LD information and nearby correlated variants (rcond = 0.01, R^2^ > 0.6, and a minimum LD threshold of 5). Fine-mapping was performed using SuSiE-RSS^9^ model on quality-controlled summary statistics, with the number of causal components (L) optimized iteratively (initial = 5, maximum = 10, increment = 5) within each LD block as described in previously published work^36^. Regions without strong signals (PIP < 0.025) are skipped automatically. Posterior inclusion probabilities (PIP) were used to define 95%, 70%, and 50% credible sets, providing fine-mapping results at multiple confidence thresholds.

Fine-mapping diagnostics were then conducted to evaluate the reliability and interpretability of credible sets, providing an automated quality assessment of fine-mapping results. For loci without identified credible sets, the variant with the highest PIP was retained. Correlation matrices were computed to quantify interdependence among credible sets by calculating the maximum and minimum absolute correlations. Each locus was subsequently assigned an optimal fine-mapping update strategy based on the number, significance, and correlation of credible sets, selecting among Bayesian Variable Selection Regression (BVSR), Single Effect Regression (SER), and Bayesian Conditional Regression (BCR) models. Following automated evaluation, results were curated through manual exploratory analysis. Posterior credible set metrics were aggregated at the study-block level, where correlation matrices between credible sets were used to detect redundant or overlapping signals (e.g., |r| > 0.8) and to assess variant sharing across studies. Genomic blocks were classified into single– or multi-credible set categories, and models with unusually large credible sets were flagged as potentially unstable. Manual inspection included visualization of PIP distributions, cross-credible set correlation structures, and concordance with marginal P-values, allowing exclusion of outlier or low-confidence regions.

### Colocalization analysis

Multi-context colocalization analyses were performed using two complementary strategies with *ColocBoost*^11^:

(1) Multi-GWAS colocalization for gene prioritization. This strategy integrated DNE traits and AD GWAS traits (**Table S1**) for joint modeling of association signals. Both DNE and AD GWAS analyses used the same quality-controlled summary statistics and the same LD reference panel from the ADSP/GCAD consortium, consistent with the fine-mapping analyses.
(2) Molecular QTL colocalization across multi-omic contexts for prioritized gene validation. Multi-GWAS colocalization was also performed across diverse molecular phenotypes, including bulk and single-nucleus expression, splicing, and protein QTLs from the ROSMAP cohort generated by the FunGen-xQTL Consortium (**Table S2**).

All analyses were performed using the ColocBoost framework. Regions with weak signals (PIP < 0.025) were excluded for splicing QTLs, with all other parameters set to default.

### TWAS analysis

Transcriptome-Wide Association Studies (TWAS) analyses were performed for xQTL molecular traits to assess their causal associations with DNE, aging, and AD GWAS traits (**Table S2**).

For each molecular trait, the genome was partitioned into LD blocks^36^, consistent with the fine-mapping analysis, and within each block, prediction models were trained for all genes whose transcription start sites (TSS) were located within the corresponding region. Six predictive models, including Elastic Net, LASSO, Bayesian Ridge, Bayesian LASSO, Sum of Single Effect (SuSiE), and Mr.ASH^34,37–41^, were trained to estimate cis-xQTL weights for candidate genes following the optimized FunGen-xQTL protocol. Elastic Net combines both L1 and L2 regularization penalties, whereas LASSO applies only the L1 norm. Bayesian Ridge and Bayesian LASSO extend linear regression by incorporating Gaussian and double-exponential priors, respectively, to model uncertainty in effect-size estimates. SuSiE captures the joint effects of multiple causal variants within a probabilistic framework, while Mr.ASH uses an empirical Bayes approach to estimate effect-size distributions across traits. Model performance was evaluated using five-fold cross-validation, and models with mean R^2^ > 0.01 were retained for downstream analyses.

After model training, the corresponding GWAS summary statistics were harmonized with the same ADSP LD reference data. Associations between genetically predicted molecular trait levels and GWAS z-scores were then tested using burden-based statistics to derive TWAS z-scores and P-values. A TWAS signal was considered significant if the best-performing model (1) had a TWAS p-value below (2.5 × 10^−6^) in the global landscape analysis, or (2) remained significant after Bonferroni correction (Bonferroni corrected p < 0.05).

### Mendelian Randomization

Fine-mapped xQTL variants with a cumulative posterior inclusion probability (cPIP) > 0.5 were used as instruments. For each molecular trait, xQTL and GWAS estimates were aligned after allele matching and standard quality control. Exposure effects were scaled by their standard errors, and credible-set level causal effects were obtained by weighting ratio estimates by their posterior inclusion probabilities. Inverse-variance weighted credible-set estimates were integrated across credible sets using fixed-effect meta-analysis to derive gene-level MR effects. Cochran’s Q and the corresponding I² statistic were used to assess heterogeneity. Gene-level MR associations were considered significant when at least one credible set was present, cPIP > 0.9, the meta-analytic p-value < 0.05, and I² < 0.5.

### Definition of xQTL-associated genes for DNE traits

Genes were prioritized from DNE-associated variants through integration with xQTL data using significant colocalization and TWAS associations, following the strategy outlined below.

(1) Colocalization: Variants supported by the multi-GWAS colocalization analysis were intersected with fine-mapped xQTL variants from the FunGen-xQTL Consortium across bulk and single-nucleus expression, splicing, proteomic, and glycoproteomic datasets. For each xQTL trait, we included variants from credible sets at multiple coverage levels (95%, 70%, and 50%), along with additional variants outside these sets that had posterior inclusion probability of at least 0.25, to broaden the set of candidate variants. Genes linked to overlapping variants were designated as prioritized targets.
(2) Putative causal gene (PCG) analysis: Merged TWAS results across multiple brain cohorts were loaded and filtered for robust models (cross-validation R^2^ > 0.01, P_cv_ < 0.05) and significant associations (P_TWAS_ < 2.5 × 10^−6^). For each LD block and molecular context, the gene containing the variant with the highest posterior inclusion probability (PIP) within the 95%, 70%, or 50% credible set was selected as the most likely causal candidate for evaluation using DNE GWAS summary statistics. GWAS summary statistics were integrated at the chromosome level, and missing p-values were recalculated from z-scores when necessary. The variant with the smallest GWAS p-value within each region was then identified as the putative causal variant, and its corresponding gene was extracted as the final prioritized target.

Genes were prioritized into three tiers based on integrated evidence from TWAS and colocalization analyses across DNE, aging, and AD traits. Tier 1 included the most confident candidates supported by both TWAS and colocalization signals in all three trait categories (AD, Aging, and DNE). Tier 2 comprised genes showing both TWAS and colocalization evidence in DNE traits but only TWAS support in AD and Aging. Tier 3 included genes with either TWAS or colocalization evidence in DNE traits and only TWAS support in AD and Aging.

### Gene-set enrichment analysis

Genes from the union of colocalization-prioritized and causal TWAS-prioritized results were identified as significant xQTL gene targets for each DNE trait and subjected to Gene Ontology (GO)^42^ and Kyoto Encyclopedia of Genes and Genomes (KEGG)^43^ enrichment analyses using the R packages *clusterprofiler*^44^ to characterize their functional relevance for each DNE trait. Gene lists were compared across DNE traits using either overlapping or union sets when applicable. GO analyses were performed across all three ontology categories: biological process (BP), cellular component (CC), and molecular function (MF). All annotated human genes in the default *clusterProfiler* background (*org.Hs.eg.db*) were used as the reference set for enrichment. Pathways with Benjamini-Hochberg adjusted p < 0.1 were considered statistically significant. Results from GO and KEGG enrichment were combined and jointly visualized to highlight functional enrichments across different DNE traits.

Genes reaching significance in both TWAS (p < 2.5 × 10⁻⁶) and Mendelian randomization were assessed by over-representation analysis using msigdbr^45^ (v25.1.1) to access curated human gene sets from MSigDB. Enrichment analyses were conducted to delineate biological processes pertinent to Alzheimer’s disease or telomere-length biology. Pathways with p < 0.05 were considered significant.

### Heritability enrichment analysis

We quantified the heritability enrichment of functional annotations using the stratified linkage disequilibrium score regression (sLDSC)^46^ framework implemented in the *polyfun* package. The sLDSC model estimates the proportion of SNP-based heritability attributable to specific functional categories by regressing GWAS □^2^ statistics on LD scores partitioned by annotation, thereby assessing whether variants within annotated regions explain more heritability than expected under a null model of uniform heritability per SNP.

The variants with maximum variant colocalization probability (maxVCP) were extracted from all Colocalized Confidence Sets (CoS) and uncolocalized CoS (UCoS) in multi-GWAS colocalization results for each DNE trait and compiled into annotation files indicating SNP membership within DNE trait-specific regions. AD and aging GWAS summary statistics were used to estimate SNP-based heritability and evaluate functional enrichment across all annotations, after filtering variants with MAF ≤ 0.001 and imputation quality ≤ 0.6, and excluding strand-ambiguous and human leukocyte antigen (HLA) region variants. Stratified LD score regression (S-LDSC) was conducted jointly with the 97-category baseline LD v2.2 model, leveraging approximately 8 million autosomal SNPs and LD scores computed from the ADSP reference panel^33^. The model estimated the marginal annotation coefficient (tau), representing the per-SNP contribution of each annotation to heritability, and the enrichment statistic, defined as the proportion of heritability explained by SNPs within a given annotation relative to their genomic coverage.

### Genetic correlation analysis

Pairwise genetic correlations between imaging-derived neurodegenerative dimensions (DNEs) were estimated using bivariate LD Score Regression (LDSC)^48,49^. We used the same genome-wide association summary statistics generated in the sLDSC processing, restricted to autosomal HapMap3 SNPs to satisfy LDSC model assumptions. Standard QC filters were applied to remove variants with missing allele information, duplicated positions, or extreme test statistics.

Bivariate LDSC was performed using LD scores and regression weights from the 1000 Genomes Project Phase 3 European (HapMap3) reference panel. For each pair of DNE traits, LDSC regressed the product of Z-scores from the two GWAS onto the HapMap3 LD scores to estimate genetic covariance and genetic correlation (rg). Pairwise rg values and standard errors were visualized in a genetic correlation matrix to summarize the polygenic similarity structure among DNE, Aging and AD traits, the Bellenguez et al. dataset was treated as a proxy AD cohort and the Kunkle et al. dataset as a clinical AD cohort.

### GTEx tissue expression analysis

Gene expression across tissues was quantified using GTEx v10 RNA-seq data downloaded from GTEx Portal (https://gtexportal.org). For each gene of interest, TPM values were extracted and merged with the corresponding tissue assignments. Tissue-level expression distributions were summarized across samples and visualized using boxplots on a log_10_ TPM scale, ordered by the mean expression across tissues.

### Pathology association analysis

Genome-wide association analyses were performed for nine harmonized neuropathological traits in ROSMAP (TDP-43 pathology, Tangles burden, Cerebral atherosclerosis, Neurofibrillary tangle burden, Cerebral amyloid angiopathy, Cognitive decline slope, Neuritic plaque burden, Amyloid burden, Arteriolosclerosis). For each trait, single-variant association testing was conducted using PLINK^50^ v1.9 under an additive linear model, adjusting for age at death, sex, and the first four genetic principal components.

### Data and Code Availability

All data supporting the findings of this study are provided in the main text and Supplementary Information. Summary statistics generated by the FunGen-xQTL project will be made available upon publication of the accompanying consortium manuscript. The complete analysis codebase, including workflows for fine-mapping, colocalization, TWAS, and functional enrichment, is available at https://github.com/rl3328/DNE_xQTL_paper.

## ACKNOWLEDGEMENTS

We thank members of the Wang, Wen, and Davatzikos labs for discussions and support. We thank the members of the Alzheimer’s Disease Sequencing Project Functional Genomics Consortium (FunGen-AD) for providing the FunGen-xQTL resource. This work was supported by NIH/NIA R01AG076901 and R01AG086467(G.W., R.L.), a FunGen-AD Consortium Grant U01AF072572 (G.W., R.F., P.L.D.), a grant from The Urbut Family Foundation (G.W.).

## AUTHOR CONTRIBUTIONS

R.L., R.F. contributed equally. G.W., J.W., C.D. jointly supervised research. R.L., R.F., J.W., G.W. conceived and designed the experiments, performed the analyses, and interpreted the results. X.C., A.L. contributed statistical methodology. P.L.D., D.B., G.W., R.F. contributed to and supervised other molecular QTL data production. C.D. and J.W. contributed to and supervised the DNE data production. R.L., R.F., G.W. wrote and revised the manuscript.

## COMPLETING INTERESTS

The authors declare no competing interests.

